# Distinct immune-effector and metabolic profile of CD8^+^ T Cells in patients with autoimmune polyarthritis induced by therapy with immune-checkpoint inhibitors

**DOI:** 10.1101/2022.03.09.22272158

**Authors:** Karolina Benesova, Franziska V. Kraus, Rui A. Carvalho, Leonore Diekmann, Janine Günther, Karel D. Klika, Petros Christopoulos, Jessica C. Hassel, Hanns-Martin Lorenz, M Margarida Souto-Carneiro

**Author notes:** **Corresponding Author:** Dr rer nat Margarida Souto-Carneiro, Division of Rheumatology, Department of Internal Medicine V, University Hospital Heidelberg, Otto-Meyerhof-Zentrum, Rm 01.123, Im Neuenheimer Feld 350, 69120 Heidelberg, Germany. KB and FVK contributed equally to this work.

## Abstract

**Objectives:** Rheumatic immune-related adverse events (irAE) such as (poly)arthritis in patients undergoing immune checkpoint inhibitor (ICI) treatment pose a major clinical challenge. ICI-therapy improves CD8+ T cell (CD8) function, but CD8 contribute to chronic inflammation in autoimmune arthritis (AA). Thus, we studied whether immune-functional and metabolic changes in CD8 explain the development of musculoskeletal irAE in ICI-treated patients.

**Methods:** Peripheral CD8 obtained from ICI-treated patients with and without musculoskeletal irAEs and from AA-patients with and without history of malignancy were stimulated in media containing ^13^C-labeled glucose with and without Tofacitinib. Changes in metabolism, immune-mediator release, expression of effector cell-surface molecules, and inhibition of tumor cell growth were quantified.

**Results:** CD8 from irAE patients showed significantly lower frequency and expression of cell-surface molecules characteristic for activation, effector-functions, homing, exhaustion and apoptosis and reduced release of cytotoxic and pro-inflammatory immune-mediators compared to CD8 from ICI-patients who did not develop irAE. This was accompanied by a lower glycolytic rate. Gene-expression analysis of pre-ICI-treated CD8 revealed over 30 differentially expressed transcripts in patients who later developed musculoskeletal irAEs. *In vitro* Tofacitinib treatment did not significantly change the immune-metabolic profile nor the capacity to inhibit the growth of the human lung-cancer cell-line H838.

**Conclusions:** Our study shows that CD8 from ICI-treated patients who develop a musculoskeletal irAE have a distinct immune-effector and metabolic profile from those that remain irAE-free. This specific irAE profile overlaps with the one observed in CD8 from AA-patients and may prove useful for novel therapeutic strategies to manage ICI-induced irAEs.

**Key messages:** 

**What is already known about this subject?:**

1. Immune-checkpoint inhibition (ICI) therapies have a high success rate regarding progression free and overall survival of cancer patients. However, up to 20% of the ICI-treated patients develop musculoskeletal immune-related adverse events (irAE) that are often associated with severely reduced quality of life.
2. To avoid precocious ICI-treatment termination, strategies to treat rheumatic irAE have to be simultaneously efficient in curbing musculoskeletal symptoms without interfering with the antitumoral therapy.
3. CD8^+^ T cells play a pivotal role both in arthritis pathogenesis and antitumoral responses.

**What does this study add?:**

1. Immuno-functional and metabolic analysis of peripheral CD8^+^ T cells from patients with musculoskeletal irAEs revealed that they share a common profile with those from patients with chronic autoimmune polyarthritis (AA) but are distinct from ICI-treated patients who remained irAE-free.
2. CD8^+^ T cells from irAE patients treated *in vitro* with the JAK-pathway inhibitor Tofacitinib still maintained the capacity to release cytokines and cytolytic molecules, express immune-effector cell surface molecules, and prevent the growth of a human lung-cancer cell line.

**How might this impact on clinical practice or future developments?:**

1. The specific immuno-functional and metabolic profile in rheumatic irAEs and its overlap to AA-profile is a potential starting point for a better understanding of the pathogenesis and identification of ICI-patients at risk of developing an irAE.
2. JAK inhibitors may expand the thus far limited therapeutic armamentarium to cope with severe, refractory and / or chronical rheumatic irAEs.

## Introduction

Immune-checkpoint inhibition (ICI) therapies that prevent CTLA-4 and PD-1 from blocking T cell activation are a milestone in cancer management. Their initial success in patients with advanced melanoma and non-small-cell lung cancer (NSCLC) has encouraged their use for other types of solid tumors[1, 2]. The increase in the number of patients under ICI-therapy is rising the number of patients developing ICI-induced immune-related adverse events (ICI-irAE) resembling chronic autoimmune diseases[3], including rheumatic musculoskeletal and systemic symptoms as well as flares of pre-existing inflammatory diseases[4]. *De novo* arthralgia, inflammatory arthritis, tendinitis/tenosynovitis, enthesitis, and (poly-)myalgia have been reported in about 20% of ICI-patients in clinical trials, with a large variation in the prevalence due to differing criteria and awareness for these side effects[4, 5]. The development of ICI-induced irAE has been associated with a better survival and clinical outcome[6-8], including patients with rheumatic irAEs[9-11]. However, severe irAEs may force clinicians to terminate ICI-therapy due to an ICI-irAE-associated mortality in 0.5-1.5% of patients[12]. Fortunately, except for myositis, rheumatic irAEs are seldom fatal but can cause considerable suffering and disability. In contrast to other ICI-irAEs, rheumatic irAEs regularly take a chronic course and require long-term medication[13]. While numerous severity-based treatment algorithms for rheumatic irAEs have been formulated to reduce inflammation and patient suffering[4, 5], there is an unmet need for evidence-based anti-inflammatory approaches without negative effects on the beneficial antitumor response in this population[9, 14-17].

In this context, data from our and other groups support the hypothesis that CD8^+^ T cells (CD8) play an important role in maintaining chronic arthritis and their permanent pro-inflammatory effector phenotype is fuelled by an enhanced aerobic glycolysis[18-20]. While the use of therapies that reduce the CD8 cytotoxic pro-inflammatory potential such as Janus-kinase inhibitors (JAKi) may be beneficial to control autoimmune arthritis (AA), they might be inappropriate for irAE, since, a fully functional CD8 anti-tumoral response is crucial for a long-term remission[21]. However, CD8 seem to play a role in the induction and/or propagation of irAE, since patients with irAE present a clonal expansion of CD8 in the periphery prior to symptom development[22] and gene-expression profiles of CD8 from irAE patients are distinct from those who do not develop irAE[8]. Nonetheless, functional studies on CD8 in irAE patients that could contribute information to evaluate this therapeutic target are largely missing. Therefore, the aim of the present study was to characterize the immuno-functional and metabolic phenotype of the peripheral CD8 pool in patients with rheumatic irAEs and compare these to the CD8-profiles from patients who did not develop irAE under ICI-treatment (ICI-CNT), patients with autoimmune arthritis (AA-CNT), and patients with autoimmune arthritis and a clinical history of malignancy (AA-MAL). We then explored JAK inhibition as a potential therapeutic strategy in ICI-irAEs and AA-MAL by testing whether *in vitro* blockade of the JAK-pathway in CD8 of these patients results in a major loss of functionality and metabolic remodelling.

## PATIENTS AND METHODS

A detailed description of the patient selection, the experimental and statistical methods can be found in the online supplementary materials file 1.

### Study approval and patient and public involvement

The study was officially approved by the institutional ethics committee (ethic votum numbers: S-096/2016, S-391/2018, S-686/2018). All subjects signed an informed written consent prior to any study procedure.

## RESULTS

### Patient characteristics

Demographic and clinical data regarding malignancy and autoimmune characteristics are summarized in table 1. Further details on underlying rheumatic diseases, irAEs, and malignancies of individual patients are listed in supplementary table 1. Most ICI-patients had a diagnosis of stage III or IV melanoma or NSCLC and all had at least a stable disease as best response. Half of the ICI-irAE patients and all in the ICI-CNT group were still under ICI-treatment at sample collection. The ICI-CNT group had a shorter disease and ICI-treatment duration and higher proportion of males. Musculoskeletal irAEs were verified and treated by a rheumatologist and were characterized by inflammatory arthralgia/arthritis, tenosynovitis and/or polymyalgia, including one patient with an overlap of polymyalgia and suspected mild myositis, another with overlap of SpA and acute gout, one with concomitant scar sarcoidosis as further irAE, and two with a flare of either pre-existing RA or a pre-existing PsA. Treatment consisted mainly of low-dosed glucocorticoids (GC)≤ 10 mg prednisolone-equivalent with only one patient receiving a higher dose at sample collection. Two patients required methotrexate and one received leflunomide for GC-sparing, none were treated with biologic (b) or targeted synthetic DMARDs. Only one patient showed high disease activity and elevated CRP at sample collection.

**Table 1.**
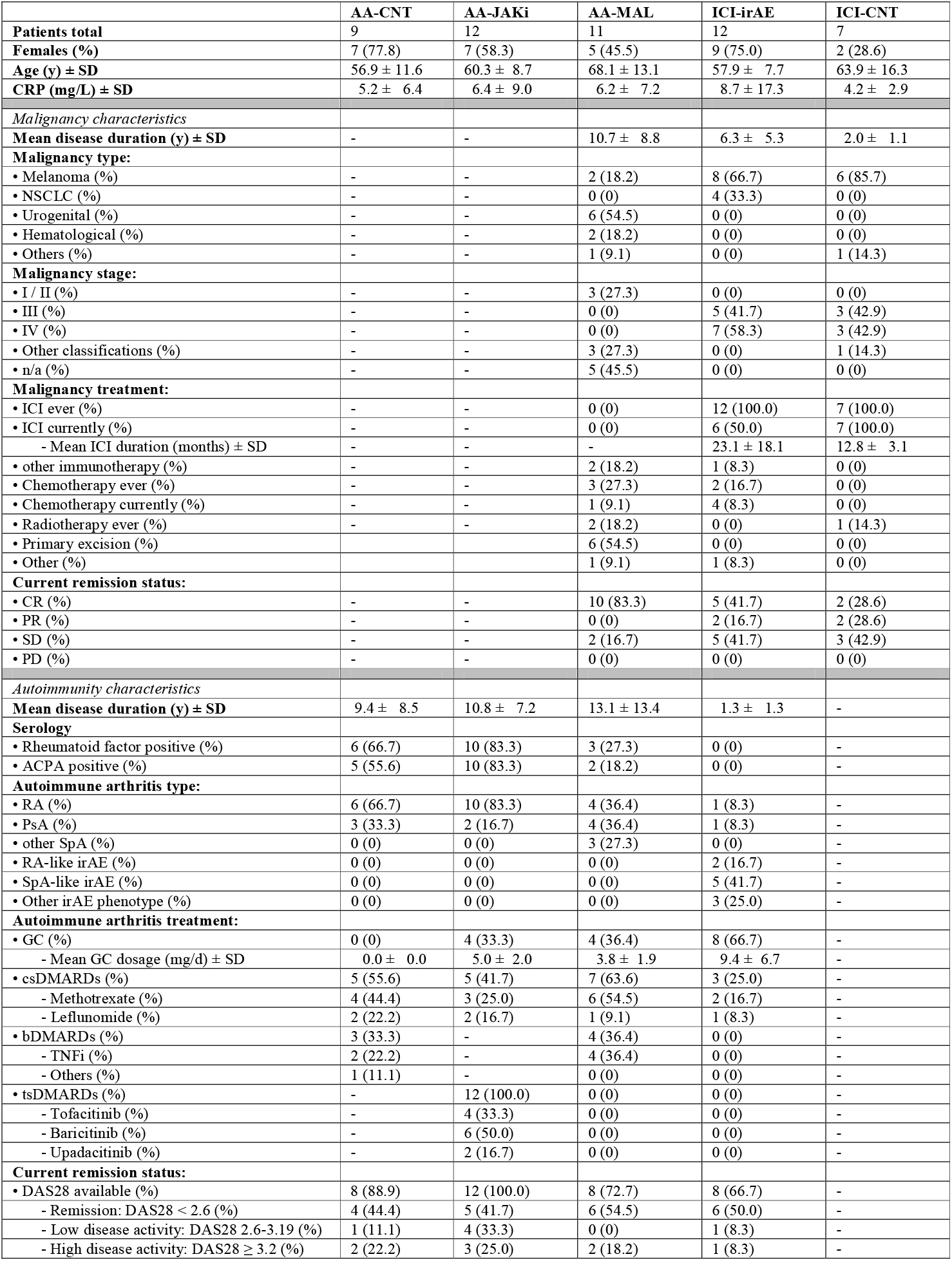
Clinical and Demographic characteristics of the study participants

The AA-MAL patients had a longer duration and a larger spectrum of malignant diseases though most patients showed complete remission. Most of the AA-MAL group received conventional synthetic (cs) and/or bDMARDs at sample collection, while GC were used less and in a lower dosage that in the ICI-irAE group. In contrast to the AA-MAL group, the AA-CNT and AA-JAK groups consisted of patients of younger age, male gender, shorter duration of the rheumatic disease, and higher rates of predominantly rheumatoid factor and/or ACPA positive RA. CRP levels were low to normal across all AA groups.

### Expression of cell-surface markers and release of immune mediators distinguishes the CD8 between patient groups

To determine expression differences in CD8 cell-surface molecules characteristic for activation and effector functions, homing, and exhaustion and apoptosis upon TCR-mediated stimulation, we projected the data onto a two-dimensional UMAP organized by the expression of the common molecules conserved among the pooled data of all study individuals followed by unbiased FlowSOM analysis for population clustering (Figure 1A-C, Supplementary Figure SF1A). The distribution of naïve, effector, effector memory and central memory subsets within the total CD8 population was similar between all study groups (χ^2^=8.66, p=0.74). Even though the expression of other cell-surface markers was considered in the FlowSOM analysis, the identified meta-clusters mirrored the division into the four CD45RA versus CCR7 subsets, and their distribution was also not significantly different (χ^2^=20.54, p=0.49; Figure 1D). Significant fold-changes from baseline in the frequency and expression of each cell-surface marker on total CD8 and in the release of cytotoxic mediators and cytokines were observed after TCR-mediated stimulation (Figure 1E-G). Differences in the expression of cell-surface molecules and soluble mediators were observed between TCR-stimulated AA-CNT and AA-MAL or ICI-irAE, and between ICI-CNT and ICI-irAE (Supplementary Figure SF1B-D). We analysed whether these differences could distinguish AA-CNT from the AA-MAL and ICI-irAE groups (Figure 1H). A higher expression of Granzyme A and PD-1 were characteristic for AA-MAL and ICI-irAE CD8 in comparison to AA-CNT. Further molecules that separated these groups from AA-CNT were the cytolytic molecules sFasL and Granulysin (AA-MAL) and CTLA-4 (ICI-irAE). We observed that ICI-irAE CD8 were distinguished from ICI-CNT by a lower expression of activation and homing molecules, pro-inflammatory cytokines (IFN-γ, TNF-α), and several cytolytic mediators. When searching for molecules that were distinct between ICI-CNT and AA-CNT, we found that they mostly overlapped with the ones separating ICI-CNT from ICI-irAE

**Figure 1.**
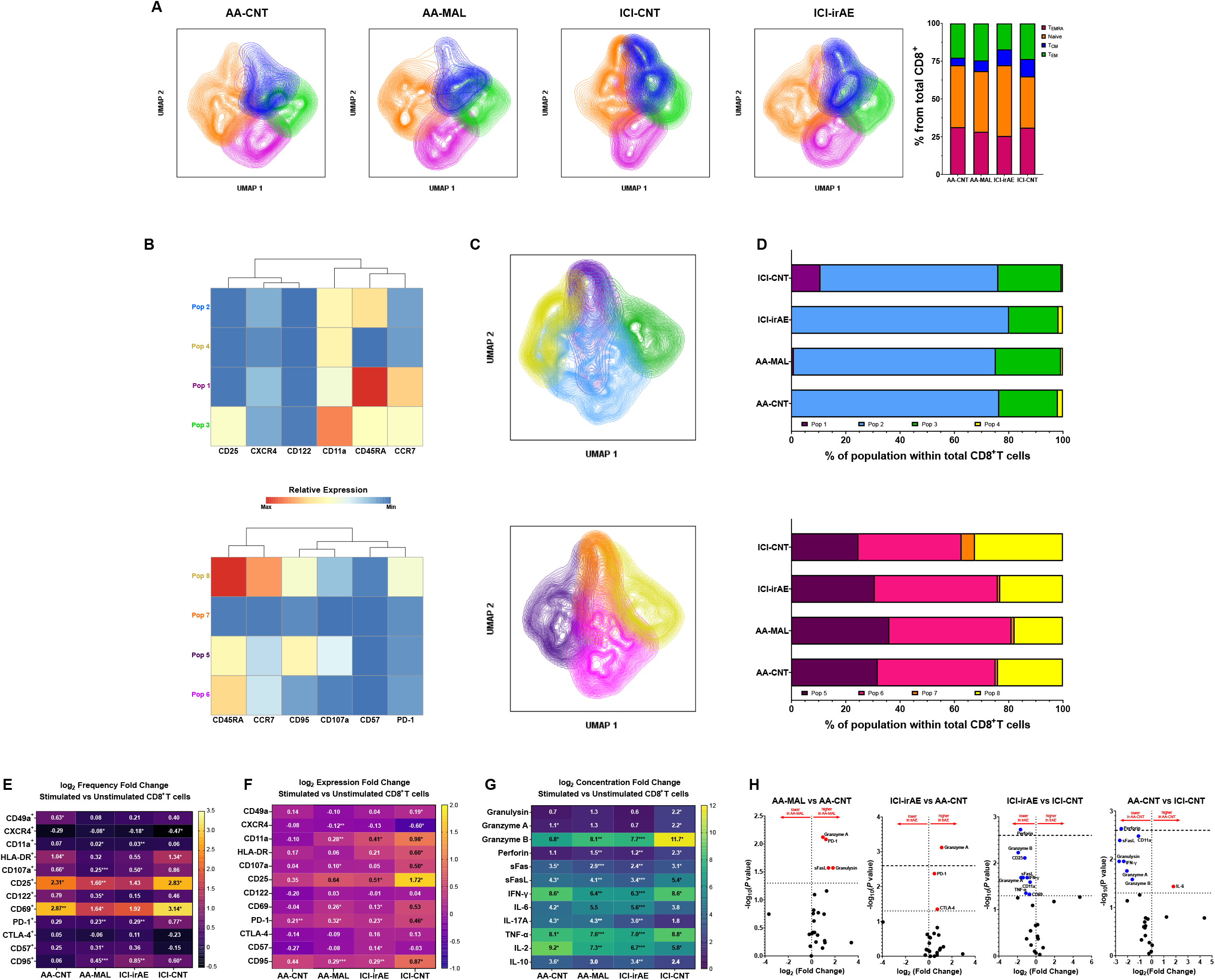
A) UMAP and stacked-column graphs showing the distribution of the four main functional CD8+ T cell subsets based on CD45RAxCCR7 expression (naïve: CD45RA+CCR7+; TEMRA: CD45RA+CCR7-; TEM: CD45RA-CCR7-; TCM: CD45RA-CCR7+) within each patient group. B) Heatmaps showing the expression (MFI) of the different markers in the meta-clusters identified by the FlowSOM analysis. C) UMAP graphs mapping the distribution of the FlowSOM meta-clusters. D) Stacked-bar graphs depicting the distribution of each meta-cluster within each patient group. E-G) Heatmaps showing the fold-change in surface-marker frequency (E), surface marker expression (F) and cytokines and cytotoxic molecules release (G) after TCR-mediated stimulation. Bold numbers: significant (p<0.05) change between stimulated and unstimulated;* p<0.05, ** p<0.01; ***p<0.001 in AA-MAL or ICI-irAE means compared to AA-CNT, in ICI-irAE means compared to ICI-CNT. H) Volcano plots showing the differentially expressed molecules between the different patient groups. Horizontal dotted line represents p-value<0.05; horizontal dashed-line represents adjusted p-value<0.05.

To determine whether clinical or demographic characteristics could contribute to and explain any of the described differences, we correlated the continuous clinical (including treatment modalities) and demographic variables with the experimental data for both the whole study cohort and for each patient group. There were no clinical or demographic variables with a significant correlation with the CD8 phenotype across all study groups (Supplementary Figure SF2).

### Metabolic phenotype of CD8

Since the cell-culture media contained [U-^13^C]-glucose, the *in vitro* glucose consumption, and *de novo* [U-^13^C]-lactate production could be precisely quantified by ^1^H-NMR (Figure 2A). CD8 from AA-CNT increased glycolysis upon *in vitro* TCR-stimulation, characterized by a strong *de novo* [U-^13^C]-lactate synthesis and a reduced rate of oxidative phosphorylation (OXPHOS). Though this trend was apparent in AA-MAL and ICI-irAE, only ICI-CNT CD8 switched to a hyper-glycolytic metabolism comparable to AA-CNT (Figure 2B-D). Additionally, we evaluated glucose consumption against expression levels of the glucose transporter GLUT1. Slight, but statistically insignificant, positive correlations were perceptible for AA-CNT and ICI-irAE (Figure 2E). The release of pro-inflammatory cytokines and cytolytic molecules positively correlated with increasing [U-^13^C]-lactate concentrations, particularly the ICI-CNT CD8 (Figure 2F). We did not find any general or group-specific correlation between clinical or demographic variables and GLUT1 expression or [U-^13^C]-lactate production (Supplementary Figure SF2).

**Figure 2.**
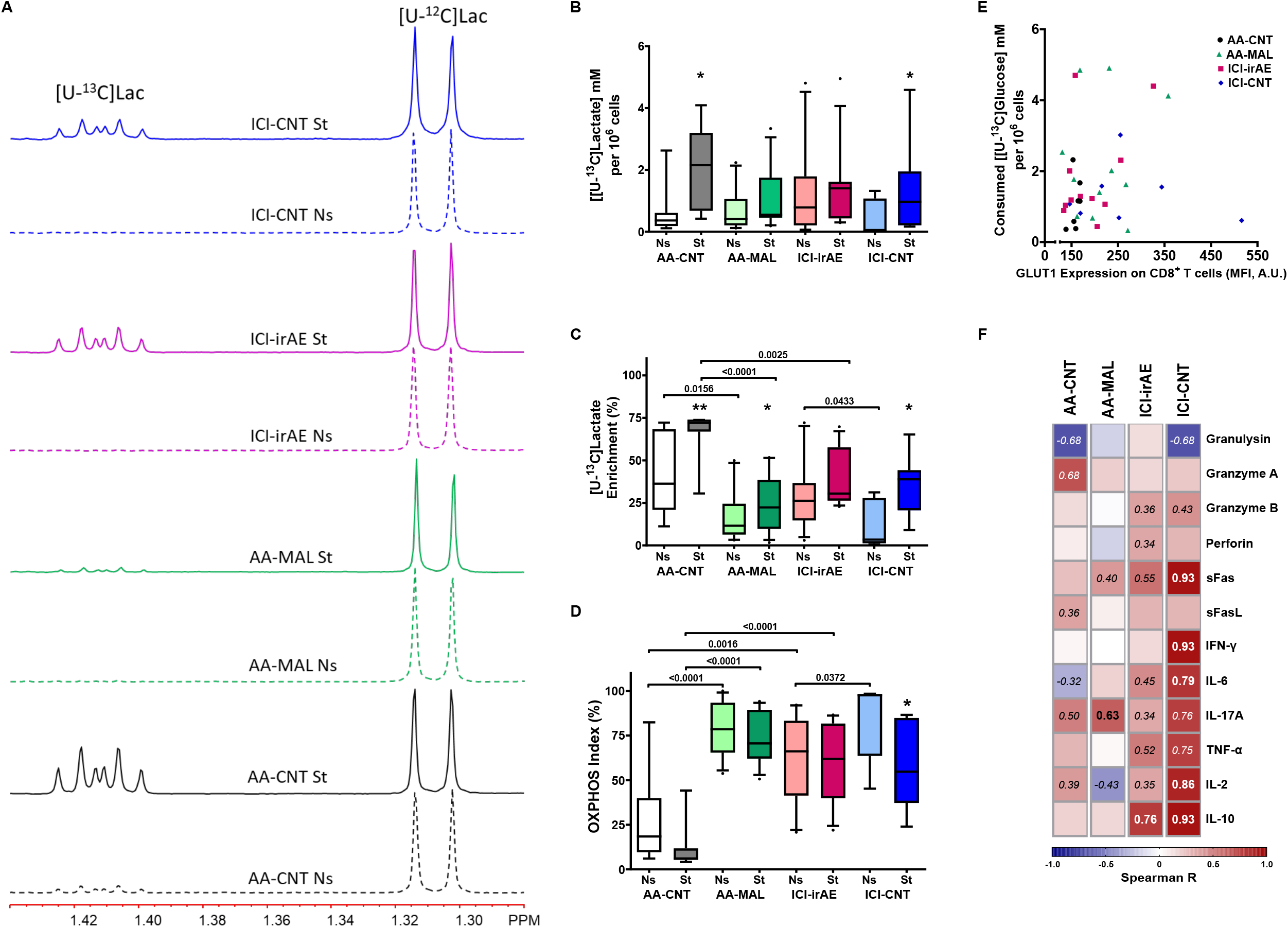
A) Representative ^1^H NMR sub-spectra of cell-culture media for each group of CD8 cells, either unstimulated or TCR-stimulated. The region covers the [U-^12^C]-lactate methyl signal and the ^13^C satellite at higher frequency arising from [U-^13^C]-lactate. Each spectrum has been normalized separately to its [U-^12^C]-lactate methyl signal. B-D) The concentration of [U-^13^C]-lactate in the cell-culture medium (B), [U-^13^C]-lactate enrichment (C), and OXPHOS-rate (D) before and after TCR–mediated stimulation for each group. Results are shown as box plots. Each box represents the 25^th^ to 75^th^ percentiles. Lines inside the boxes represent the median. Lines outside the boxes represent the 10^th^ and 90^th^ percentiles. Circles represent outliers. Asterisks represent significant * p<0.05, ** p<0.01; ***p<0.001 changes between stimulated and unstimulated conditions. E) X-Y plot showing the correlation between [U-^13^C]-glucose uptake and GLUT1 expression after TCR-mediated stimulation. F) The correlation between [U-^13^C]-lactate production and cytokines/cytotoxic molecules release upon TCR-mediated stimulation. Numbers show correlations with Spearman R >|0.3|, bold numbers represent p<0.05.

### Different baseline gene-expression profiles distinguish CD8 from ICI-patients who develop musculoskeletal irAE

We retrieved the EGAS00001004081 gene expression data[8] obtained from peripheral CD8 isolated before the beginning of ICI-therapy and compared the profiles of patients who later developed arthritis irAE (13.5%) with those who did not develop any ICI-induced irAE. 22 transcripts were differentially expressed between the whole group of patients who developed musculoskeletal irAE and those who did not, and pathway analysis revealed an enrichment of genes involved in cell population proliferation, immune system development, and response to TNF in the group that remained irAE-free (Figure 3A; Supplementary Table ST3). Before ICI-treatment with only anti-PD-1, we identified 47 transcripts and an enrichment of pathways linked to ATP metabolism and immune response which were differentially expressed in those patients who later developed musculoskeletal irAE (Figure 3B; Supplementary Table ST4). No major gene-expression differences were observed within the patients who received a combination of anti-CTLA-4 and anti-PD-1 (Supplementary Table ST5). Based on arthritis severity, we identified 18 transcripts that were differentially expressed, but we could not define any significantly altered pathway. Data did not indicate which patients would require GC to curb musculoskeletal irAE (Supplementary Table ST6). All these data indicate that CD8 differ even before the initiation of ICI.

**Figure 3.**
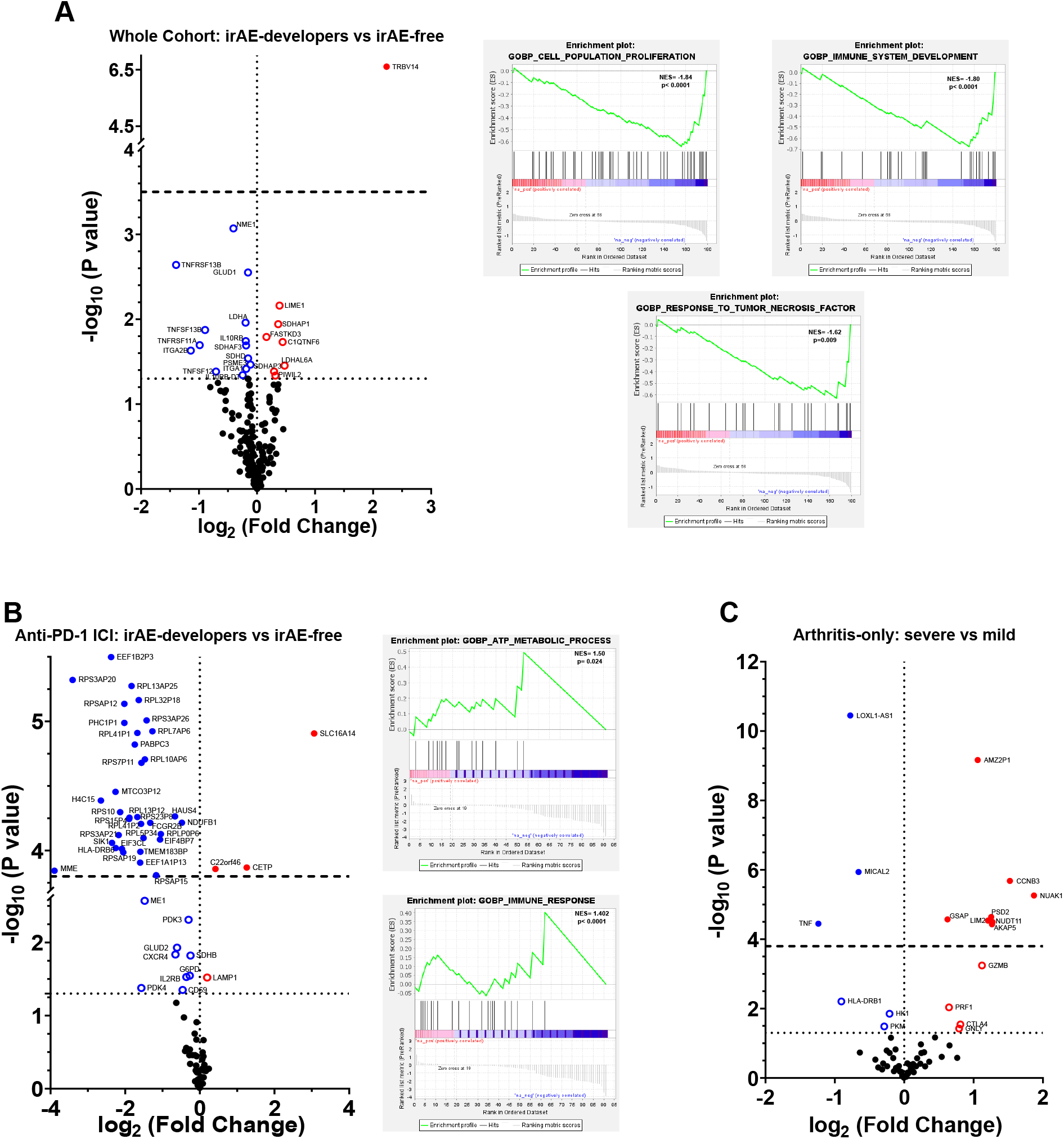
Volcano plots and pathway enrichment plots showing the gene expression differences before ICI-therapy. A) Total ICI-treated patients. Developed arthritis irAE (n=21) versus remained irAE-free (n=135). B) Patients treated only with anti-PD-1 ICI. Developed arthritis irAE (n=7) versus remained irAE-free (n=73). C) ICI-treated irAE patients who developed severe arthritis (grade 3-4; n=7) versus those who developed mild arthritis (grade 1-2; n=14). Horizontal dotted line represents p<0.05; horizontal dashed-line represents adjusted p<0.05.

### *In vitro* inhibition of JAK-signaling pathway inhibition does not induce major functional or metabolic changes in CD8

To evaluate the effect of JAKi, TCR-stimulated CD8 of all groups were compared to *in vitro* JAKi-treated CD8. Additionally, TCR-stimulated AA-CNT CD8 were compared to *in vitro* TCR-stimulated CD8 from AA patients under *in vivo* JAKi therapy (AA-JAK group).

JAKi led to a decrease in the frequency of CD8 expressing activation markers on all groups. While AA-JAK patients had less CD8 expressing homing markers than AA-CNT, *in vitro* JAKi increased the frequency of CD8^+^CXCR4^+^ cells for AA-MAL, ICI-CNT, and ICI-irAE and had no impact on the frequency of CD8^+^CD11a^+^ and CD8^+^CD49a^+^ after stimulation. The frequency of CD8 displaying exhaustion markers was generally lower after JAKi (Figure 4B). A similar pattern was observed for the cell-surface expression of these molecules (Figure 4A and C). The differences observed when comparing TCR-stimulated AA-CNT CD8 to those from AA-JAK patients were like those observed in TCR-stimulated AA-CNT CD8 under *in vitro* JAKi treatment (Supplementary Figure SF3A-B). Even though *in vitro* JAKi-treatment led to a generalized reduction in the concentration of soluble mediators, only ICI-CNT CD8 released significantly less pro-inflammatory and cytolytic mediators upon *in vitro* JAKi treatment (Figure 4D).

**Figure 4.**
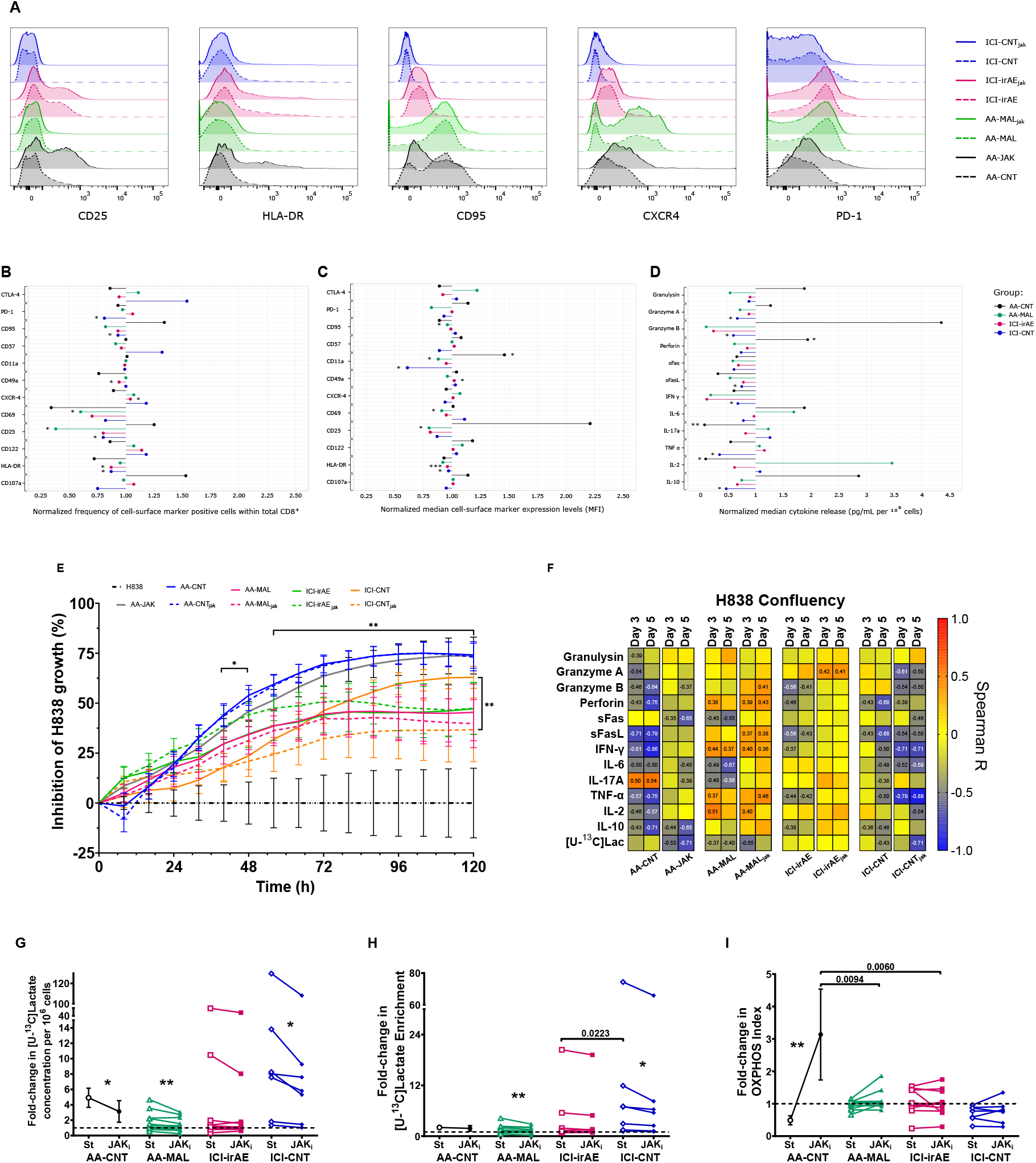
A) Representative histograms of changes in the expression of cell-surface molecules by TCR-stimulated CD8+ T cells after in vitro JAKitreatment (AA-MAL, ICI-irAE, and ICI-CNT) or between AA-CNT and AA-JAK patients. B-D) Lollipop graphs showing the fold-changes in surface-marker frequency (B), surface marker expression (C), and cytokines and cytotoxic molecules release (D) of TCR-stimulated CD8+ T cells after *in vitro* JAKi treatment (AA-MAL, ICI-irAE, and ICI-CNT) or between AA-CNT and AA-JAK patients. * p<0.05, ** p<0.01; ***p<0.001 changes between stimulated and JAKi conditions. E) Inhibition of H838 growth by conditioned media from CD8+ T cells (TCR-stimulated with and without JAKi treatment). Two-way ANOVA: horizontal * p<0.05, ** p<0.01 between media from all patient groups versus H838 in unconditioned medium for the indicated timepoints; vertical ** p<0.01 between JAKi-treated and untreated ICI-CNT. F) Correlations between H838 cell growth and the concentration of cytokines or cytotoxic molecules in the conditioned cell-culture media. Numbers show correlations with Spearman R >|0.35| and p<0.05. G-I) Fold-change relative to baseline in the concentration of [U-^13^C]-lactate in the cell-culture medium (G), [U-^13^C]-lactate enrichment (H) and OXPHOS rate (I) in TCR-stimulated CD8+ T cells with (solid symbols) or without (open symbols) *in vitro* JAKi treatment (AA-MAL, ICI-irAE, and ICI-CNT) or between AA-CNT and AA-JAK patients. * p<0.05, ** p<0.01 between JAKi-treated and untreated cells.

The *in vitro* expansion of H838 cells could be inhibited for 5 days using conditioned-medium from TCR-stimulated CD8 (Figure 4E; Supplementary Figure SF3C-D). the conditioned-media from AA-CNT and AA-JAK patients’ CD8 were the more inhibitory. Except for ICI-CNT, conditioned-media from CD8 cultured in the presence of JAKi had comparable inhibitory capacity on H838 cell-growth than media without JAKi. Consistent significant negative correlations between H838 inhibition and the concentration of cytokines or cytolytic molecules were evident in the conditioned-media from CD8 from AA-CNT, AA-JAK and ICI-CNT patients (Figure 4F).

The metabolic profile of CD8 was altered upon *in vitro* JAKi treatment. We observed less [U-^13^C]-lactate accompanied by more OXPHOS when comparing AA-CNT to AA-JAK CD8. Less [U-^13^C]-lactate production and enrichment, along with a slightly higher, yet not significant OXPHOS-rate, could also be observed for *in vitro* JAKi-treated AA-MAL and ICI-CNT CD8 (Figure 4G-I; Supplementary Figure SF3E).

## Discussion

Functional and phenotypical changes in CD8 have been generally associated with the success of antitumor response and are thus the core of ICI-therapy[23, 24]. However, such changes are equally contributing to the pathophysiology of chronic AA[19, 20, 25]. This poses a major challenge in the treatment of ICI-induced arthritis as inhibition of CD8 would be required for sustained arthritis therapy which would, however, limit the antitumor effects. To clarify some of these aspects, we have compared phenotype and functional and metabolic changes in peripheral CD8 from ICI-patients who either developed or did not develop musculoskeletal irAEs with those from AA patients and tested the effects of *in vitro* JAKi on the antitumor functions of CD8. The identified effector CD8 meta-clusters were slightly more abundant in ICI-irAE, AA-CNT and AA-MAL than in ICI-CNT patients. This enrichment of effector/activated CD8 in ICI-irAE patients has been equally been reported in the peripheral blood of thymic epithelial tumor and metastatic NSCLC patients developing different forms of irAEs[26], in the epidermis of melanoma, renal cell carcinoma, gastric cancer and lung cancer patients with ICI-induced psoriasis-like dermatitis[27], and in the colon epithelium of melanoma patients with ICI-induced colitis[28]. Thus, they seem to help fuelling the (autoimmune) inflammatory response in ICI. Contrasting with this accumulation of effector meta-clusters, ICI-irAE CD8 -and from the other arthritis groups, AA-CNT and AA-MAL-released less pro-inflammatory and cytotoxic mediators than ICI-CNT.

Metabolic remodelling from OXPHOS towards aerobic glycolysis is a hallmark of CD8 activation[29-31], and in patients with chronic AA they display a permanent and exacerbated glycolytic profile that maintains chronic cytotoxicity[19]. Thus, it was not surprising that TCR-mediated stimulation induced aerobic glycolysis in CD8 from all ICI-patients, even though activation-induced change in the OXPHOS-rate occurred only in ICI-CNT. Since resting ICI-irAE CD8 were more glycolytic than ICI-CNT, but released less cytotoxic and pro-inflammatory mediators, it is possible that ICI-irAE CD8 have a better energetic and biosynthetic balance to sustain their effector/antitumor functions for a longer period when compared to ICI-CNT cells and might contribute to the better clinical outcomes in ICI-irAE patients. However, keeping a steady pro-inflammatory and cytotoxic effector phenotype for longer periods has the downside of rendering ICI-irAE CD8 with a RA-like profile, which favours the surge and relapse of irAE.

Gene-expression analysis has shown that the development of different irAEs has been associated with pre-and post-ICI downregulation of CXCR1 on peripheral CD8 in melanoma patients[8]. Here we re-analysed the same pre-ICI gene-expression dataset focusing on those patients who developed musculoskeletal irAEs. Even though the number of available samples was limited - which limits data interpretation - the results suggest a baseline impairment in the upregulation of TNF-signalling and proliferation pathways. These differences appear to remain after musculoskeletal irAE has developed, since CD8 from ICI-irAE patients released less TNF and expressed less CD25 than those from ICI-CNT. Collectively, these are relevant findings in the context of the lively discussion on beneficial or detrimental effects of TNF inhibition as a treatment option for ICI-patients with severe irAEs[32-34].

Currently, therapeutic algorithms for irAE-arthritis rely on defensive use of GC, csDMARDs, and TNF- or IL-6-blockers[4, 5]. However, the use of TNF-inhibition in irAEs is increasingly controversial[32-34] and our data suggest that CD8 from ICI-irAE patients present a downregulation of the TNF-signalling pathway and release less TNF than ICI-CNT. Thus, finding other therapeutic strategies to curb ICI-induced musculoskeletal inflammation and maintain antitumor activity will be required to meet current clinical needs in the management of ICI-irAEs. Preclinical studies on human cancer cell lines have shown that JAK-pathway inhibition impairs tumor growth[35, 36]. Still, new data on increased risk of malignancy in RA patients associated with Tofacitinib use put the previously favourable assessment of JAKi *in vitro* and *in vivo* into question[37, 38]. However, considering the limited treatment options, one needs to expand the therapeutic armamentarium to cope with severe and/or chronical ICI-irAEs. Therefore, the beneficial potential risks of JAKi in musculoskeletal and other irAEs should be further investigated, particularly when keeping in mind the increasing number of available JAKi with minor, but clinically relevant, differences in their modes of action. In view of this, we carried out *in vitro* experiments to explore the feasibility of using JAK-pathway inhibition by Tofacitinib to control CD8 pro-inflammatory activity without severely compromising antitumor response. Our data suggest that *in vitro* Tofacitinib did not significantly reduce the release of cytotoxic mediators by ICI-irAE CD8. Although we could only indirectly measure the *in vitro* capacity of JAKi-treated CD8 to inhibit tumor cell-growth, it seems that in our experimental setting the antitumor capacity of CD8 from ICI-patients could be maintained, even if it was lower than observed for the cancer-free AA-patients. Additionally, *in vitro* JAKi treatment maintained a high OXPHOS rate in ICI-patients CD8 without drastically reducing aerobic glycolysis, essential for maintaining antitumor functions in CD8[39].

The lack of a group of AA-free cancer patients with ongoing tumor activity and without ICI-therapy and the use of only one type of JAK-inhibitor for the *in vitro* studies (a constraint imposed by the reduced number of cells obtained from each patient) are potential limitations of our study. To counter this limitation, we included the patients AA-MAL who had simultaneously a clinical history of malignancy (some still with active tumors) and chronic arthritis, and AA-JAK patients with chronic autoimmune arthritis receiving different types and doses of JAK-inhibitors. Since the AA-MAL and the ICI-irAE CD8 presented similar profiles, even in their response to *in vitro* JAKi, we assume that ICI, other cancer therapies, or ongoing tumor activity did not play a major role in the observed immune and metabolic profile changes. Since the CD8 phenotype was quite consistent amongst all AA-JAK patients, we considered that limiting the *in vitro* studies to one type of JAKi does not reduce the meaning of our findings.

Overall, our study shows that CD8 from cancer patients who develop musculoskeletal irAEs during ICI-treatment have a distinct immune effector and metabolic profile from those ICI-patients that remain irAE free. This irAE profile is characterized by lower cytotoxic and pro-inflammatory activity and more aerobic glycolysis and overlaps with the profile observed in AA-CNT and AA-MAL CD8. This suggests that (chronic) inflammatory arthritis has a unique fingerprint that can be used to direct new therapeutic strategies for managing ICI-induced irAE. One such therapeutic approach may involve JAK-pathway inhibition that does not interfere with the antitumoral capacity of CD8 in our experimental model. Thus, future trials on tumor-bearing mice with (poly)arthritis or controlled clinical trials on ICI-irAE patients using JAKi should be the next step to improve therapeutic outcomes while maintaining ICI-efficacy together with simultaneous irAE control.

## Data Availability

All data produced in the present work are contained in the manuscript and any raw data produced in the present study are available upon reasonable request to the authors.

## Funding Disclaimer

This work was funded by the unrestricted investigator-initiated grant WI245418 from Pfizer Pharma GmbH. The funding body did not have any influence on the study design, nor on the data acquisition and analysis and interpretation. FV Kraus was supported by the Add-On fellowship of the Joachim Herz-Foundation. K Benesova: COI: Consultancy and/or speaker fees and/or travel reimbursements: Abbvie, Bristol Myers Squibb (BMS), Gilead/Galapagos, Janssen, Merck Sharp &Dohme (MSD), Mundipharma, Novartis, Pfizer, Roche, Viatris, UCB. Scientific support: Medical Faculty of University of Heidelberg, Rheumaliga Baden-Württemberg e.V., AbbVie, Novartis.

## Acknowledgements

Benjamin Fairfax and Christine Ye (Department of Oncology, University of Oxford) for granting access to the gene-expression data and help with its analysis; Holger Lorenz (Imaging Facility, Zentrum für Molekulare Biologie, University of Heidelberg) for technical aid with the IncuCyte microscopy data acquisition and analysis; the physicians and nursing teams of the skin-cancer ambulatory unit of the National Center for Tumor Diseases, Heidelberg University Hospital; and the nursing team of the blood collection unit of the Medical Clinic, Heidelberg University Hospital.

## Notes

### Author Declarations

The study was officially approved by the institutional ethics committee of the University of Heidelberg (ethic votum numbers: S-096/2016, S-391/2018, S-686/2018). All subjects signed an informed written consent prior to any study procedure.

