## Supplementary figures and images for "Distinct immune-effector and metabolic profile of CD8^+^ T Cells in patients with autoimmune polyarthritis induced by therapy with immune-checkpoint inhibitors"

### Supplemental Figure 1

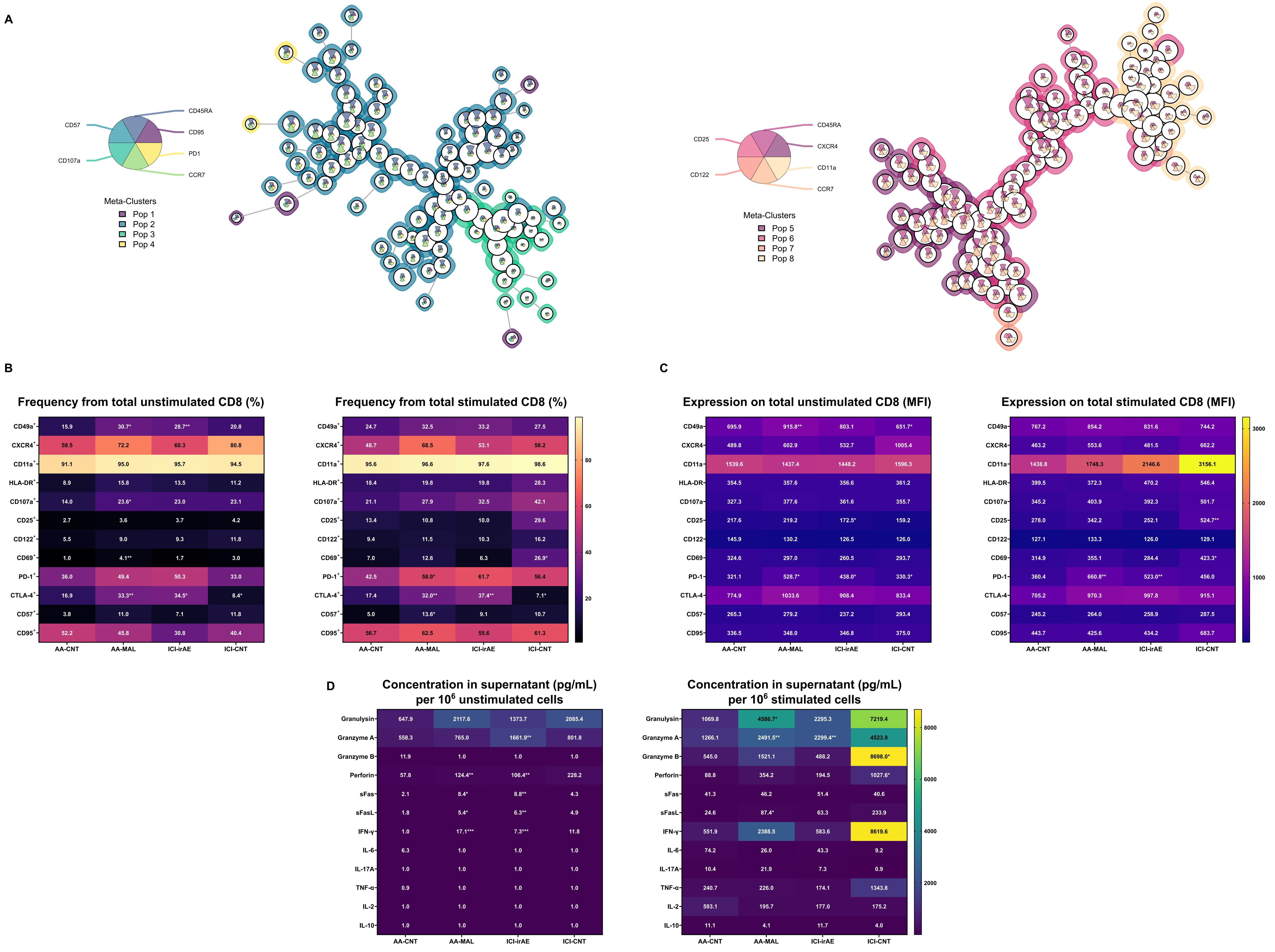

### Supplemental Figure 2

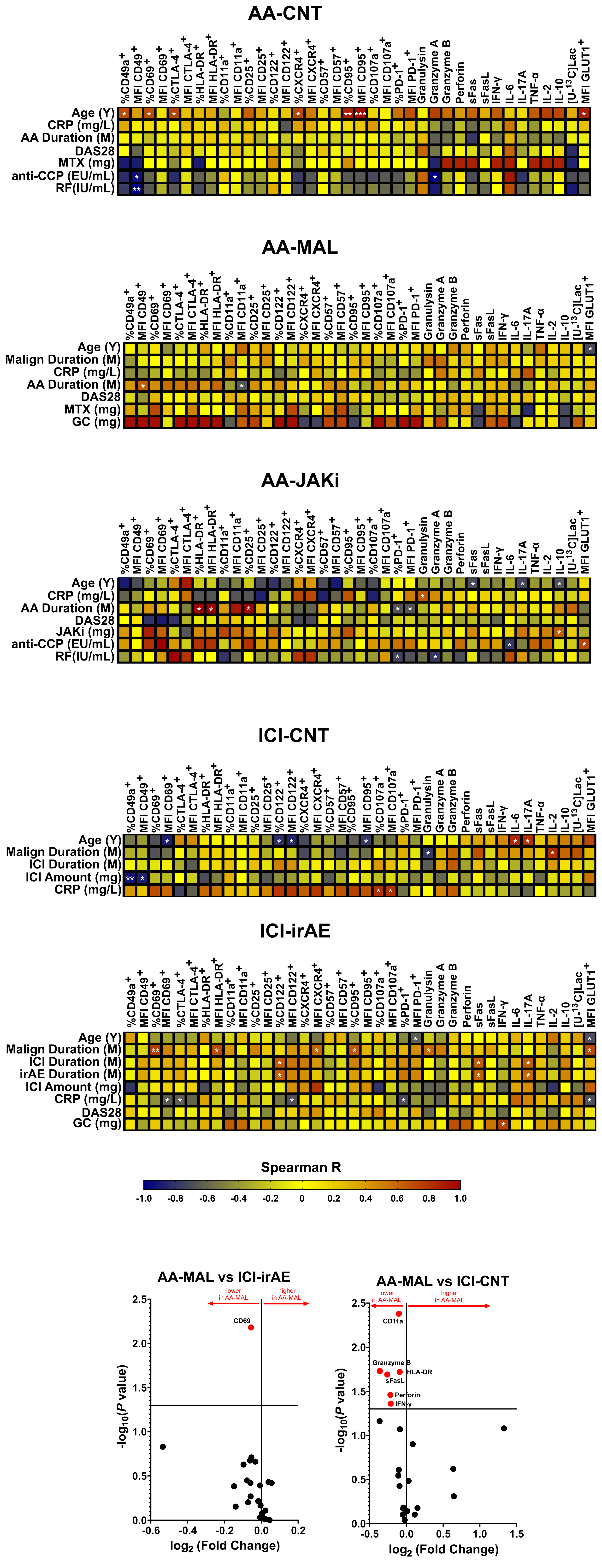

### Supplemental Figure 3

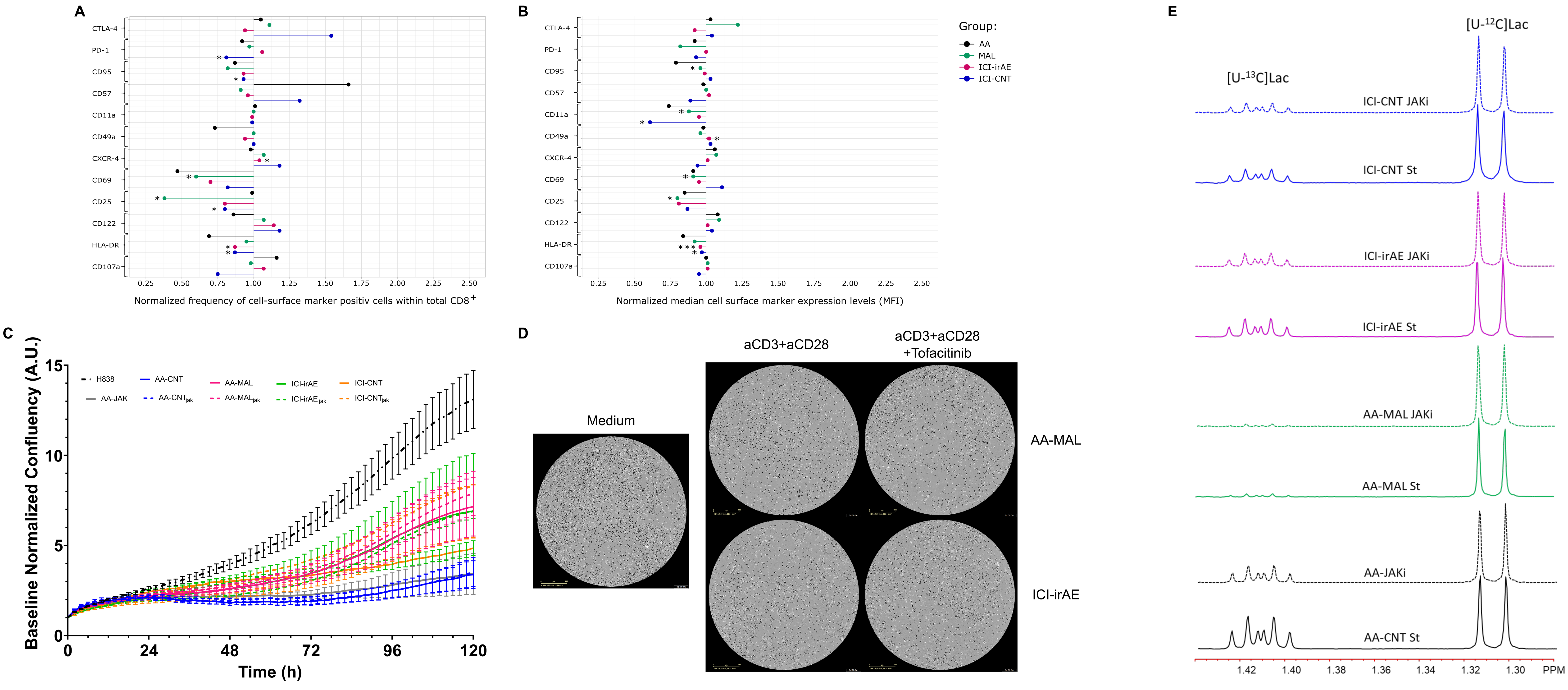
