## Supplemental Materials and Methods for "Distinct immune-effector and metabolic profile of CD8^+^ T Cells in patients with autoimmune polyarthritis induced by therapy with immune-checkpoint inhibitors"

**Supplementary Materials and Methods:**

**Patient recruitment and sample collection**

Autoimmune arthritis (AA) patients included in the study suffered from either rheumatoid arthritis (RA, by 2010 ACR/EULAR classification criteria [1], psoriatic arthritis (PsA, by CASPAR criteria [2] or other spondyloarthritis subtypes (SpA, by ASAS criteria [3, 4]. Heparinized blood samples were collected from 6 RA and 3 PsA patients under disease-modifying anti-rheumatic drugs (DMARDs) (AA-CNT group), 10 RA and 2 PsA patients under JAKi therapy (AA-JAK group), and 11 AA-MAL patients with RA, PsA, or SpA without history of ICI treatment at our Division of Rheumatology outpatient clinic. AA-CNT and AA-JAK patients were excluded from the study whenever any of the following criteria were met: 1) daily corticosteroid intake >10 mg; 2) presence of type I or II diabetes; 3) without history of malignancy other than non-melanotic skin cancer; 4) pregnancy; or 5) active viral and/or bacterial infection.

Furthermore, samples from 12 ICI-patients with musculoskeletal irAEs diagnosed by a rheumatologist (ICI-irAE group) as well as samples from 7 patients without ICI-induced irAE (ICI-CNT) were obtained at the National Center for Tumor Diseases.

**CD8^+^ T cell isolation and *in vitro* stimulation**

After density gradient isolation of peripheral blood mononuclear cells, CD8 were purified by immunomagnetic negative selection using the MojoSort™ Human CD8 T Cell Isolation Kit (Biolegend) with the aid of LS-separation columns (Miltenyi Biotech) according to the manufacturer’s instructions. Freshly purified CD8 were cultured *in vitro* for 72 h at 37°C under an atmosphere of 5% CO_2_ and 21% O_2_ in RPMI-1640 medium (Sigma-Aldrich) containing 5 mM [U-^13^C]-glucose (Sigma-Aldrich), 10 mM HEPES (PAA, USA), 10% heat-inactivated FCS (GIBCO), 100 U/mL penicillin, and 100 ng/mL streptomycin (both from GIBCO), either in the absence (non-stimulated, Nstim) or presence (stimulated, Stim) of anti-human CD28 (clone 28.1, 1 µg/mL, Biolegend) and plate-bound anti-human CD3 (clone OKT3, 2.5 µg/mL, Biolegend) in the presence or absence of the JAKi Tofacitnib (Pfizer) dissolved in deuterated DMSO (Eurisotop) at a concentration of 10 µM [5].

**Flow cytometric analysis of total CD8 and their subsets**

Harvested cells were stained with fluorescence-conjugated monoclonal mouse anti-human antibodies against CD3, CD8, CCR7 (CD197), CD45RA, PD-1 (CD279), CD57, CD49a, HLA-DR, CD25, CTLA-4 (CD152), CD69, CD11a, CD122, CD107a, CXCR4 (CD184), CD95 (all from Biolegend) and GLUT1 (R&D Systems) as previously described [6]. Irrelevant directly conjugated murine IgG1 and IgG2 (Biolegend) were used to control for background staining. After calibration with CST beads, single fluorochrome-stained cells were used for instrument compensation and PMT setup. Samples were run on an LSR II flow cytometer (Becton Dickinson) with a minimum of 5000 events collected within the CD3^+^CD8^+^ gate after doublet exclusion. Data were quantified using FlowJo Software (Treestar). The four major CD8^+^ T cell subsets were defined according to the combined expression of CD45RA and CCR7, namely naïve (CD45RA^+^CCR7^+^), effector (CD45RA^+^CCR7^-^), central memory (CD45RA^-^CCR7^+^) and effector memory (CD45RA^-^CCR7^-^). Quantification of the expression of each cell-surface marker was determined using the median fluorescence intensity for the cells positive for the marker. UMAP and FlowSOM analysis were carried out after down-sampling using the corresponding FlowJO plugins. Parameters for UMAP v3.1: 2 components, Euclidean distance calculation, 15 nearest neighbors, and 0.2 minimum distance. Parameters for FlowSOM v2.9: 4 meta clusters, 100% node scale, and 73 seed.

Cell-surface markers were divided into 3 functional classes: activation and effector functions (CD69, CD25, CD122, CD107a, and HLA-DR), homing (CXCR4, CD49a, and CD11a), and exhaustion and apoptosis (PD-1, CD57, CTLA-4, and CD95).

**Quantification of cytokine and cytotoxic molecules production by CD8**

The titers of cytokines (IL-10, IFN-γ, IL-6, TNF-α, IL-2, and IL-17A), and cytotoxic molecules (Granzyme A, Granzyme B, Granulysin, soluble Fas, soluble Fas-Ligand, and Perforin 1) present in the media of CD8^+^ T cell cultures were determined using the NK/CD8 LEGENDPlex assay panel (Biolegend) according to the manufacturer’s instructions. CBA raw data analysis was performed using the LEGENDplex v8.0 software (Biolegend) and concentrations were corrected for the number of cells.

**NMR analysis of cell-culture media**

The ^1^H NMR spectra of cell-culture media were acquired using a 600 MHz Bruker spectrometer equipped with a 5-mm indirect-detection probe as previously described [6]. Spectral analyses were performed using NUTSpro^TM^ NMR software (Acorn NMR Inc., Livermore, CA, USA). Sodium fumarate (10 mM) in 0.2 M phosphate buffer prepared with D_2_O (99.9%) was used as internal standard for quantification. NMR samples of cell-culture media consisted of 160 μL of culture media plus 40 μL of fumarate standard. The ^13^C satellite signals of lactate in the ^1^H spectrum derived from the metabolism of isotopically labeled [U-^13^C]-glucose were used for calculating the concentration of lactate for the evaluation of glycolysis and oxidative metabolism. Metabolite concentrations were assessed by deconvolution of the ^1^H NMR spectra using the line-fitting sub-routine of NUTSpro^TM^. Metabolite concentrations in cell-culture media are reported in µmol per million cells.

**Gene-Expression analysis**

Pre-treatment CD8 transcriptomic data were obtained from an ICI-patient cohort from the University of Oxford (n=156) which included 21 ICI-treated patients who developed musculoskeletal irAE and 135 matched ICI-treated patients without irAE from the European Genome–phenome Archive under accession no. EGAS00001004081 [7]. Cohort details are summarized in supplementary table 2. Differential expression analysis between groups was performed using DESeq2 [8] on a list of pre-selected genes which included those coding for metabolic enzymes involved in major metabolic pathways, the ones coding for the cell-surface markers analysed by flow-cytometry, and those coding for cytokines and cytolytic molecules. For comparisons within the patient groups receiving single-therapy (only PD-1 blockers), combined therapy (CTLA-4 and PD-1 blockers) and between patients with mild (grade 1 or 2) or severe (grade 3 or 4) arthritis, the test design included control for the influence of age and gender. A false discovery rate (FDR) threshold of 5% was applied. Enrichment pathway analysis on fold-change ranked gene lists was performed using the GSEA v4.2.0 software [9, 10] on the gene ontology biological processes dataset with 1000 permutations and weighted enrichment statistics. Gene-set size filters: min=15 and max=500, and meandiv normalized.

**H838 cell growth in conditioned CD8 medium**

The human non-small-cell lung cancer epithelial cell line H838 was maintained in high glucose DMEM containing 10% heat-inactivated FCS, 100 U/mL penicillin, and 100 ng/mL streptomycin (all from GIBCO). Immediately prior to the growth-inhibition assays, H838 cells were stained with 0.33 µM Incucyte® Cytolight Rapid Green Dye (Sartorius AG) in 1 mL PBS for 15 min at 37°C after which the reaction was quenched with 5 mL heat-inactivated FCS. Using 10000 cells per well in a 96-well plate, H838 cells were cultured at 37°C in 5% (v/v) CO_2_ in an Incucyte S3 instrument for 5 days in either 75% fresh DMEM medium containing 25% CD8+ T cell-conditioned RPMI medium produced by CD8 from the different patient groups or 75% fresh DMEM medium containing 25% RPMI medium with 3 mM glucose (control group). The instrument was configured to whole-well mode and images (phase and green) were acquired every 2 hours. Image data were analyzed with the vendor-supplied Incucyte S3 software (Incucyte version 2019B) using the Basic Analysis module to extract confluence data from the phase channel (analysis mask parameters available upon request). Data were corrected to zero time.

**Statistical analysis**

According to the D’Agostino–Pearson omnibus normality test, none of the continuous data variables followed a normal distribution. Significant outliers were identified for each data set using the Grubb’s test/extreme studentized deviate test with an alpha value of 0.05 and those above the critical z-value were removed from the subsequent statistical analysis. Unpaired log_2_-transformed data sets were analyzed using multiple Mann-Whitney tests followed by the Holm-Šídák method to adjust the p-value for the multiple comparisons to determine the significant fold-changes between stimulated and unstimulated cells from each group for each cell-surface marker and immune mediator. Ordinary two-way ANOVA, including the row/column interaction followed by the Benjamini-Hochberg false discovery rate method (FDR<10%) to correct for multiple comparisons, was used to determine whether the frequency of all CD8^+^ T cell subsets, H838 cell-growth inhibition, the expression of all cell-surface markers, or the release of all immune mediators were significantly different between patient groups. Correlations between clinical and experimental data sets were determined using the non-parametric Spearman R. Correlations were considered moderate for |0.3|<R<|0.5|; strong for |0.5|. The Chi-Square test was used to compute the differences in the distribution of the CD45RA versus CCR7 functional subsets. Differences were considered statistically different for p<0.05.

**Supplementary Figures Legends**

**SF1-** A) Minimum spanning trees representing the FlowSOM meta clusters.

B-D) Heatmaps showing the surface marker frequency among total CD8+ T cells (B), MFI of surface markers on marker-positive CD8+ T cells (C) and concentration of cytokines and cytotoxic molecules in the cell-culture medium (D) with and without TCR-mediated stimulation. * p<0.05, ** p<0.01; ***p<0.001 in AA-MAL or ICI-irAE means compared to AA-CNT, in ICI-CNT means compared to ICI-irAE.

**SF2-** A) Correlations between experimental data and clinical and demographic parameters for each group. * p<0.05, ** p<0.01; ***p<0.001 significant Spearman R.

B) Volcano plots showing the differentially expressed molecules between the different patient groups.

**SF3-** A-B) Lollipop graphs showing the fold-changes in cell-surface marker frequency (A) and cell-surface marker expression (B) of TCR-stimulated CD8+ T cells after *in vitro* JAKi treatment in all groups.

C) Zero timepoint normalized confluences of H838 cells growing in medium alone or in CD8+ T cell-conditioned medium measured every 2h for a total 5 day period.

D) Representative whole-well IncuCyte images taken at 72h showing H838 cells growing in medium alone (medium) or cell-conditioned medium from TCR-stimulated CD8+ T cells with (aCD3 + aCD28 + Tofacitinib) or without (aCD3 + aCD8) JAKi treatment from AA-MAL and ICI-irAE patients.

E) Representative ^1^H NMR sub-spectra of cell-culture media for TCR-stimulated CD8 cells, either with or without JAKi treatment. The region covers the [U-^12^C]-lactate methyl signal and the ^13^C satellite at higher frequency arising from [U-^13^C]-lactate. Each spectrum has been normalized separately to its [U-^12^C]-lactate methyl signal. For *in vitro* JAKi treatment of AA-MAL, ICI-irAE, ICI-CNT, and AA, the spectra depict AA-CNT versus AA-JAK patients.
